# A multi-centre cross-sectional study on hepatitis B vaccination coverage and associated factors among personnel working in health facilities in Kumasi, Ghana

**DOI:** 10.1101/2024.04.30.24306647

**Authors:** Daniel Kobina Okwan, Godfred Yawson Scott, Pius Takyi, Clinton Owusu Boateng, Philemon Boasiako Antwi, Akwasi Amponsah Abrampah, Michael Agyemang Obeng

## Abstract

As part of efforts to reach the elimination target by 2030, the WHO and CDC recommend that all HCWs adhere to the 3-dose hepatitis B vaccination schedule to protect themselves against the infection. This study assessed Hepatitis B vaccination coverage and associated factors among personnel working in health facilities in Kumasi, Ghana.

A cross-sectional study involving 530 HCWs was conducted in four hospitals in Kumasi from September to November, 2023. An investigator-administered questionnaire was employed in gathering participant demographics and other information related to vaccination coverage. IBM SPSS version 26.0 and GraphPad prism 8.0 were used for analysing the data.

Even though, majority (70.6%) reported having taken at least one dose of the vaccine, only 43.6% were fully vaccinated (≥ 3 doses). More than a quarter (29.4%) had not taken any dose of the HBV vaccine. Close to a quarter (23.6%) had not screened or tested for HBV infection in their lifetime. The Statistically significant variables influencing vaccination status were age, marital status, profession and status in the hospital. Majority (44.9%) of the participants who have not taken the vaccine reported they do not have reason for not taking the vaccine and high proportion (80.1%) were willing to take the vaccine when given for free.

To combat the low hepatitis B vaccination coverage among healthcare workers in Kumasi, Ghana, amidst the significant public health threat of HBV infection, comprehensive measures are necessary. These include implementing infection prevention control programmes, enhancing occupational health and safety, and conducting health promotion campaigns in healthcare facilities. Extending and intensifying hepatitis B screening and vaccination initiatives to tertiary institutions and encouraging employers, supervisors or team leaders to provide these services nationwide are also recommended.

## Introduction

As a major risk to public health across the globe, chronic hepatitis B viral infection results in significant liver-related morbidity and mortality [1]. The hepatitis B virus (HBV) is a double-stranded DNA virus belonging to the Hepadnaviridae family [2]. An estimated 2 billion individuals worldwide apparently have been exposed to the HBV, and approximately 3 million of those cases have chronic infections that put them at risk of severe illness and even death [3,4]. Updated estimates from the World Health Organization (WHO) and the Global Burden of Disease research indicate that each year, viral hepatitis causes about 1.34 million fatalities [5]. Viral hepatitis has become the seventh biggest cause of death globally, with a 63% increase in mortality since 1990 [6]. The increased rate of HBV infection and its fatalities made WHO adopt a global hepatitis strategy in May, 2016, to eliminate viral hepatitis as a public health threat by 2030 [7]. Currently, there is no cure for HBV infection; therefore, the only way to protect people all over the world is through the hepatitis B vaccination. The HBV vaccine was discovered in the year 1982. Since untreated HBV can cause cirrhosis and hepatocellular carcinoma, this vaccination was the first of its kind to prevent the cancer [8]. When administered according to the recommended schedule, the HBV vaccine provides 90–100% of healthy newborns, children, and adults with a protective concentration of anti-HBs (≥10 mIU/mL) [9].

Africa has the largest proportion of HBV-positive people worldwide, making up 68% of the total burden [5]. Ghana, a country in Sub-Saharan Africa, has a significant public health issue with HBV illness that requires immediate attention [10]. The prevalence of chronic HBV infection in Ghana was 12.92%, according to a 2013 systematic review of data published between 1965 and 2013 [11]. According to systematic review and meta-analysis published, the prevalence of HBV infection of in Ghana was as high as 12.3% in 2016 [12] and 8.36% in 2020 [10]. Ghana has achieved progress in lowering HBV disease-related death and morbidity by introducing the pentavalent vaccine in 2002, which combines the hepatitis B, DPT, and Hib vaccines [13]. Even though the disease declined after the pentavalent vaccine was introduced, there are still issues with the vaccination that can be linked to a variety of circumstances, such as ignorance of and attitudes towards the vaccination. Adult vaccination cost has been implicated to be the cause, along with ignorance, deprivation, and resistance to change [14].

Numerous types of human contacts, including sexual and non-sexual kinds, needle-sharing and work- and health-related interactions, have been linked to HBV transmission [15]. As a result, those who work in specific occupations stand a higher chance of getting HBV infection than others. Healthcare workers (HCWs) are frequently exposed to bloodborne pathogens, which puts them at high risk of contracting HBV [14,16]. When providing care for patients who are HBV-positive, workplace exposure to the virus might happen from contaminated hospital surfaces or unintentional needlestick injuries (NSIs). According to WHO estimates, 2 million HCWs have needlestick injuries each year, and 3.3% of those individuals go on to acquire hepatitis B after suffering sharp injury [17]. Therefore, WHO and Centre for Disease Control and Prevention (CDC) recommend that HCWs get the hepatitis B vaccination as a routine preventative measure [7,18]. HCWs are the ones who will help educate and encourage the hepatitis B vaccination among the general public, thus their understanding of the virus, the infection sequelae, and the need for vaccination is very crucial in controlling the HBV infection. A study conducted by Demsiss et al. [19] indicated a high seroprevalence but poor practice of hepatitis B and C virus infection among medicine and health science students despite their good knowledge of the occupational risk of viral hepatitis infection. These students are supposed to follow good practices for hepatitis B virus infection since they will be taking over the health system as practitioners in the near future soon to come.

A study conducted among HCWs in Bantama, Ghana, highlighted unsatisfactory or poor knowledge, attitude, and practice toward the hepatitis B virus and some important aspects of viral hepatitis [15]. Another study conducted by Obeng et al. [20] found the prevalence of hepatitis B viral (HBV) infection among vaccinated HCWs to be 2.4%. This study highlights the need for all HCWs not only to complete their hepatitis B vaccination but also to do the post-vaccination serological testing to confirm immunity [20]. The study proved that HCWs who have not taken the hepatitis B vaccine stand a very high risk of contracting the infection. There is not much data on hepatitis B vaccination coverage among HCWs in the country. As Ghana is highly endemic to HBV infection, this data will be needed to ascertain if the existing control programmes are effective in achieving national targets as set by WHO for 2030 HBV elimination [7]. Therefore, this study aimed at determining the vaccination coverage and throw more light on factors influencing the vaccination uptake among personnel working in select health facilities in Kumasi, Ghana.

## Materials and Methods

### Study Design and population

A cross-sectional study was conducted in four hospitals in Kumasi from 5^th^ September to 9^th^ November, 2023. These study sites included Ashanti Regional Hospital, Suntreso Government Hospital, Maternal and Child Health Hospital and Manhyia District Hospital.

Kumasi, the second largest city in Ghana, is situated between latitudes 6.35°N and 6.40°N and longitudes 1.3°W and 1.35°W. Covering an area of approximately 150 square kilometres, it resides within the rainforest region of West Africa. The city has a population of around 2 million inhabitants [21].

In the suburbs of Kumasi [22], reported the prevalence levels of HBsAg to be 6.78% in Garrison, 9.02% in Aboabo, and 10.0% in Tafo. The overall prevalence of HBsAg sero-positivity within the study population was calculated to be 8.68%. These findings indicate that local prevalence rates of HBsAg can vary significantly within different areas of Kumasi.

A total of 530 participants were recruited using a purposive sampling technique from the four different health facilities. The sample size was calculated using the *Raosoft* sample size calculator [23]. The minimum sample size required for this study was 377 participants at 95% confidence level, 5% margin of error, and a response distribution of 50%. It was increased to 530 in order to increase the statistical power of prediction.

### Ethical consideration

Ethical approval was sought from the Committee on Human Research, Publication and Ethics (CHRPE), School of Medical Sciences, Kwame Nkrumah University of Science and Technology (KNUST) (Reference number: CHRPE/AP/331/23). Approval letters were obtained from all four study sites before the commencement of the study. All participants gave their written informed consent after the aim and procedure of the study had been explained to them.

### Inclusion and exclusion criteria

The study participants were HCWs comprising clinicians, pharmacists, laboratory scientists, nurses, midwives, administrative staff, securities and cleaners. Healthcare workers in the four study sites during the study period who were willing to participate in the study were included. Healthcare workers on fieldwork, maternity, annual, or sick leave who were unable to remain in the study area during the data-collecting period were excluded.

### Data Collection

A well-structured questionnaire was validated and administered to the participants within the framework of the study variables consisting of participant demographics and other information related to vaccination coverage. The data were collected using an investigator-administered questionnaire in a language that they could easily comprehend (English and Twi).

We engaged in pilot interviews and discussions with a cohort of healthcare workers to enhance the clarity and applicability of our research questionnaire. Their feedback enabled us to fine-tune the questionnaire, ensuring it was more accessible to patients and effectively captured pertinent information. Before commencing data collection, all researchers involved underwent training. At the conclusion of each day’s data gathering, a team of investigators meticulously reviewed the obtained data for inconsistencies and omissions. Subsequently, data cleaning procedures were implemented to ensure accuracy, consistency, and completeness of variables. Any incomplete participant responses were identified, rejected, and excluded from the dataset before analysis.

### Definition of key concepts

**HBV vaccination schedule** is the hepatitis B vaccine injection that is generally given intramuscularly in the arm (deltoid muscle) as a three-dose series on a 0,1, and 6-month schedule.

**Health care workers** in this study, include all those who work in the hospital settings were considered as healthcare workers (HCWs). They included clinicians, nursing and midwifery staff members, laboratory staff members, laboratory students, administration staff members, pharmacists, cleaners and security personnel.

**Complete HBV vaccination** refers to a participant who has taken 3 or more doses of the hepatitis B vaccine.

**Incomplete HBV vaccination** in the study, referred to all who had received only one dose or two doses of the HBV vaccine.

**Vaccination status** refers to whether or not a participant has taken any dose of the hepatitis B vaccine. Participants who have taken at least one dose of the vaccine represent “Yes” and those who have not taken any dose represent “No” vaccination status.

### Data analysis

Data entry was done using Microsoft Excel 2019 and analysis was performed using IBM SPSS Version 26.0 and GraphPad Prism version 8.0. Categorical data were presented as frequency (proportion). Multivariate logistic regression analysis was performed to evaluate the factors that influence vaccination coverage of the study participants. All statistical results obtained were considered at a significant value of p < 0.05.

## Results

### Hepatitis B vaccination status

Majority (70.6%) of the study participants had taken at least one dose of the Hepatitis B vaccine. Among those who had taken the vaccine, 43.6% were fully vaccinated (≥ 3 doses) while (27.0%) had taken one or two doses of the vaccine. Meanwhile, 29.4% indicated that they had not taken any dose of the hepatitis B vaccine (**Fig 1**).

**Fig 1:**
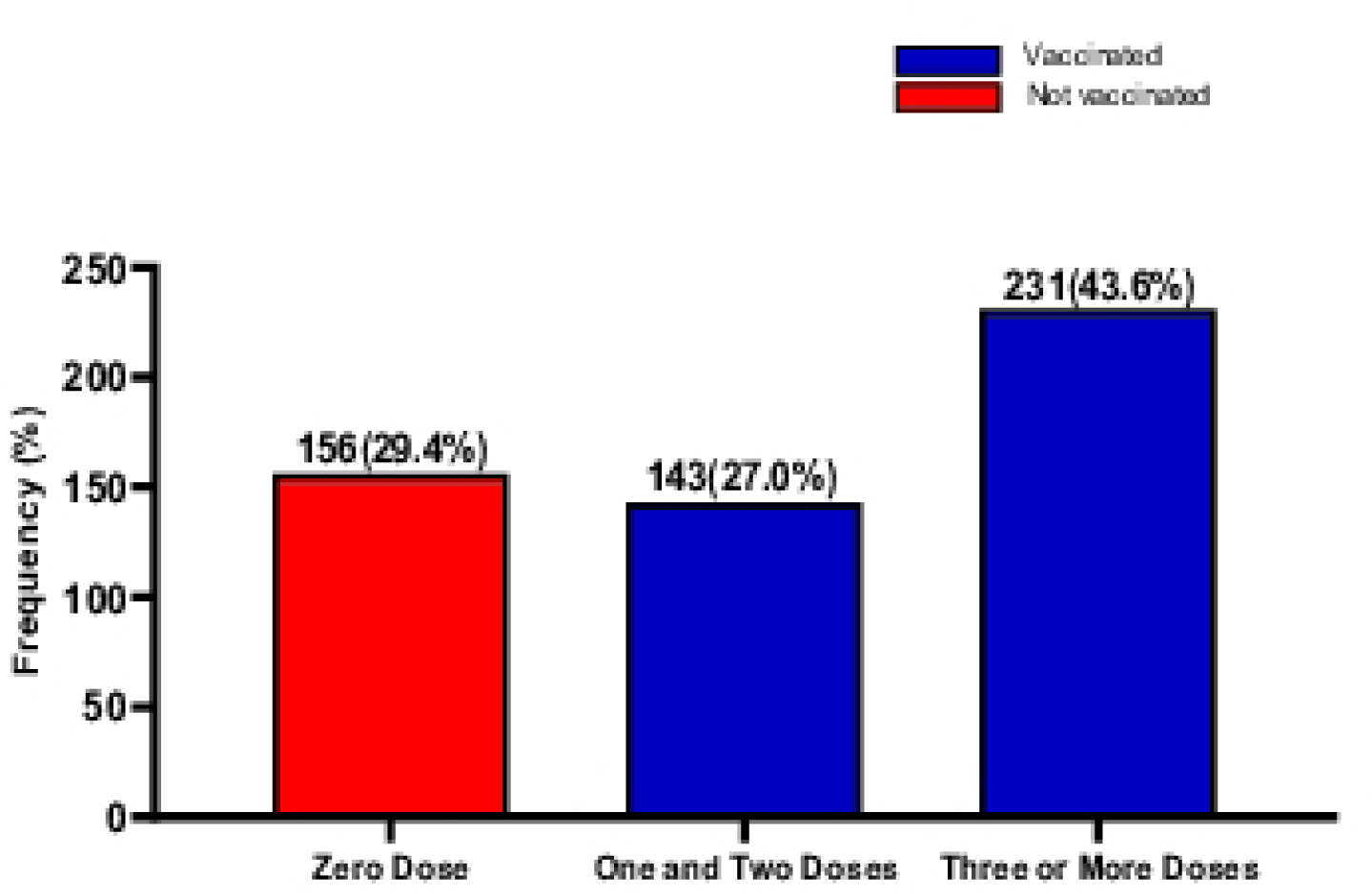
Hepatitis B vaccination status.

### Association between sociodemographic characteristics and vaccination status

Majority (66.2%) of the participants were aged 20-30 years. Most (72.6%) of them were females and were single (72.5%). Most of them were from the Ashanti region (66.6%) and were Christians (91.1%). Most of them were Diploma holders (56.6%) and were clinical staff members (83.4%) at the respective hospitals. Majority of them were contract staff members (66.2%) and were mainly from the Manhyia Government Hospital (39.4%). Individually, age group, marital status, status in hospital and profession were significantly associated with vaccination status of the participants (*p* ˂ 0.001) (**Table 1.0**)

**Table 1:**
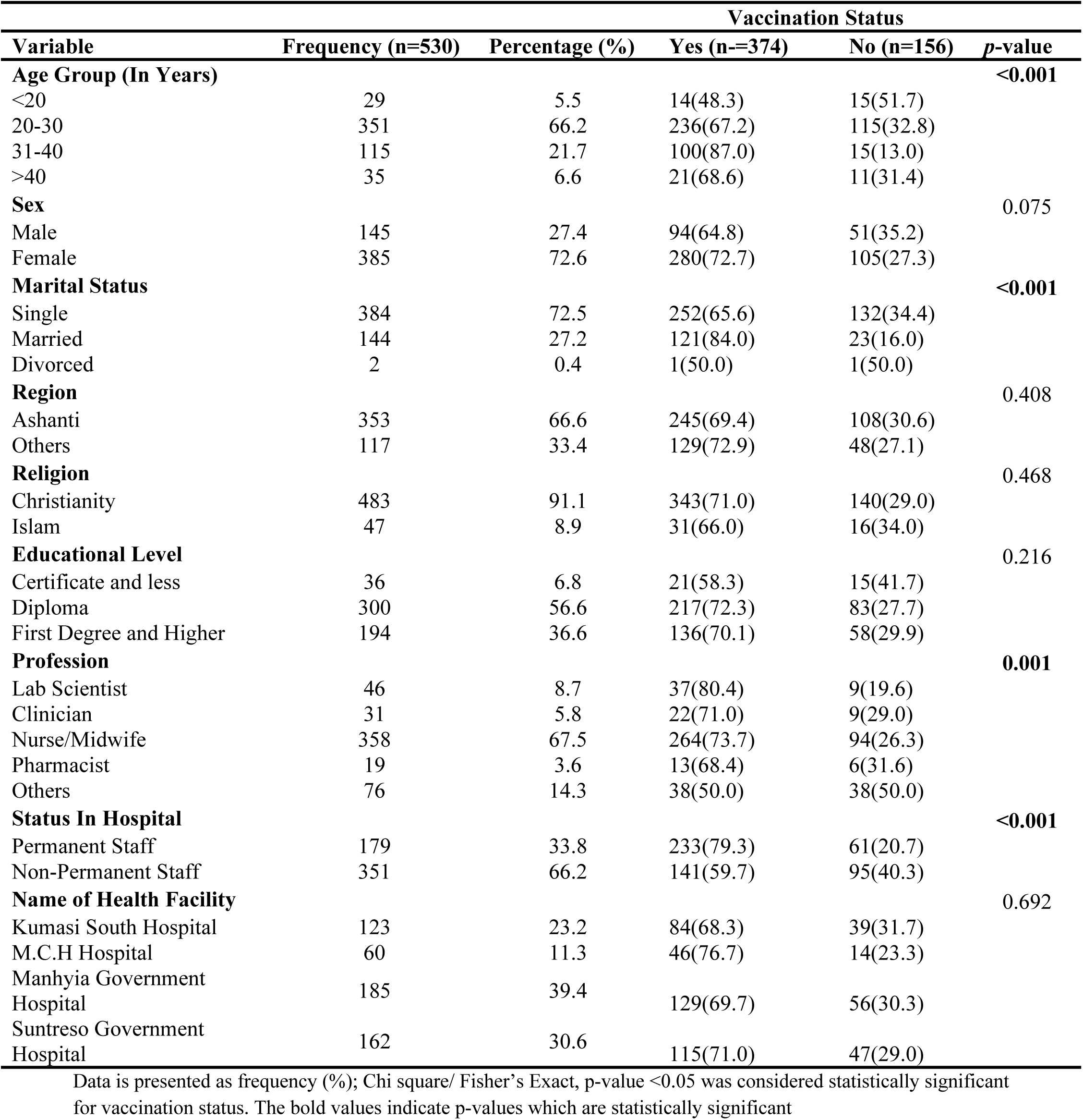
Association between sociodemographic characteristics and vaccination status.

### Association between knowledge on Hepatitis B and vaccination and vaccination status

Most (76.4%) of the participants had done the Hepatitis B test at least once in their life time, while (23.6%) had not done Hepatitis B test before. Majority (44.9) of those who had done the test reported they wanted to know their Hepatitis B status as reason for testing. Most (49.6%) of those who had not done the Hepatitis B test indicated that they did not have any reason for not testing. Also, majority (44.9%) of those who had not been vaccinated indicated that they did not have any reason for not vaccinating. Most (80.1%) of those who had not taken the vaccine were willing to take the vaccine when given to them free of charge. About (61.8%) of those who had been vaccinated had taken ≥3 doses and self-initiative (43.9%) was their source of Hepatitis B vaccination. Most (30.4%) of them indicated that the cost for Hepatitis B vaccination and testing was reasonable. Having done the Hepatitis B test before, reason for doing the test, number of Hepatitis B vaccine doses taken if vaccinated, source of hepatitis B vaccination if vaccinated, how expensive is Hepatitis B testing and vaccination, willingness to take Hepatitis B vaccine if given for free were all significantly associated with vaccination status (*p* ˂ 0.001) (**Table 2.0**).

**Table 2:**
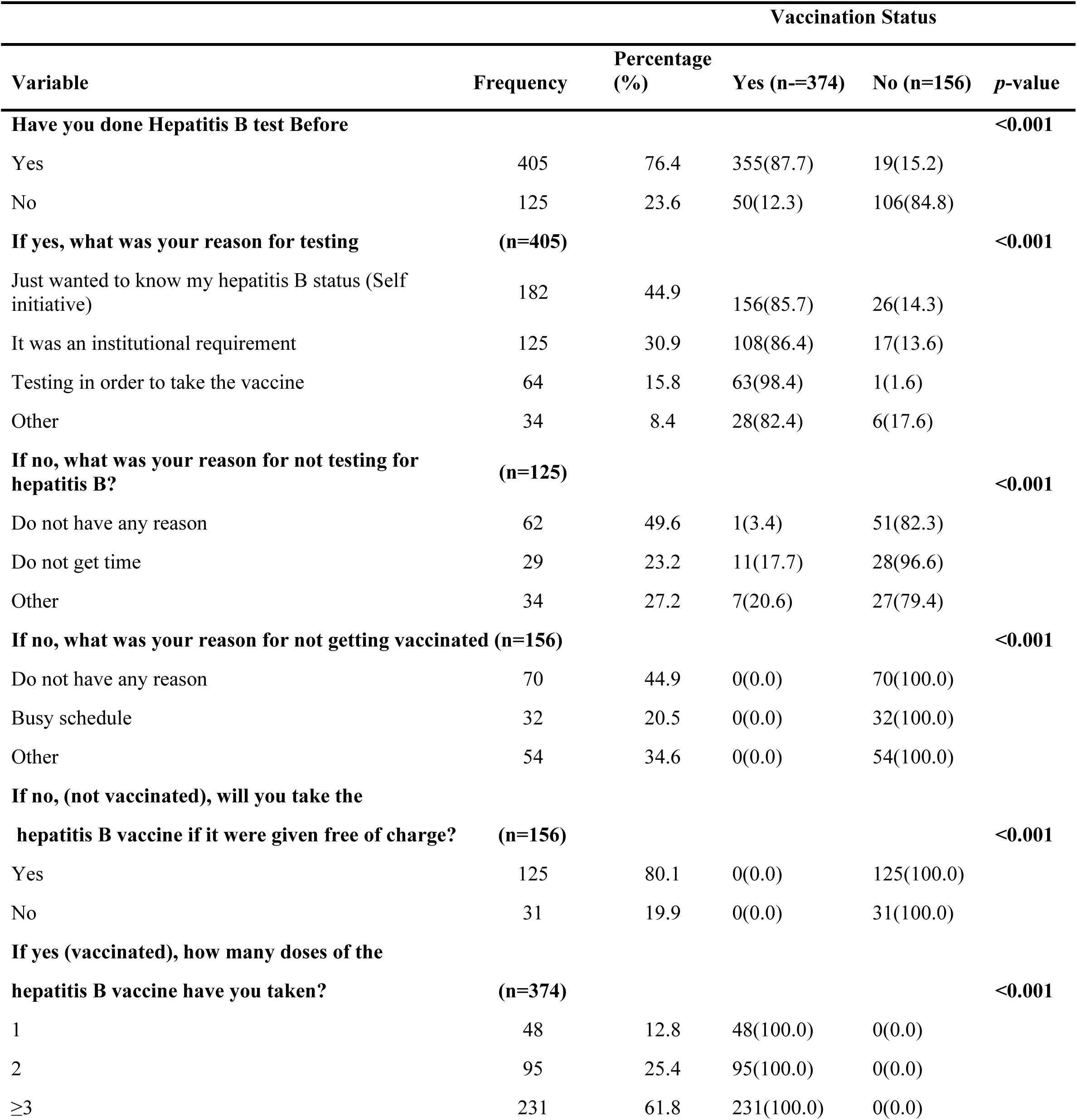
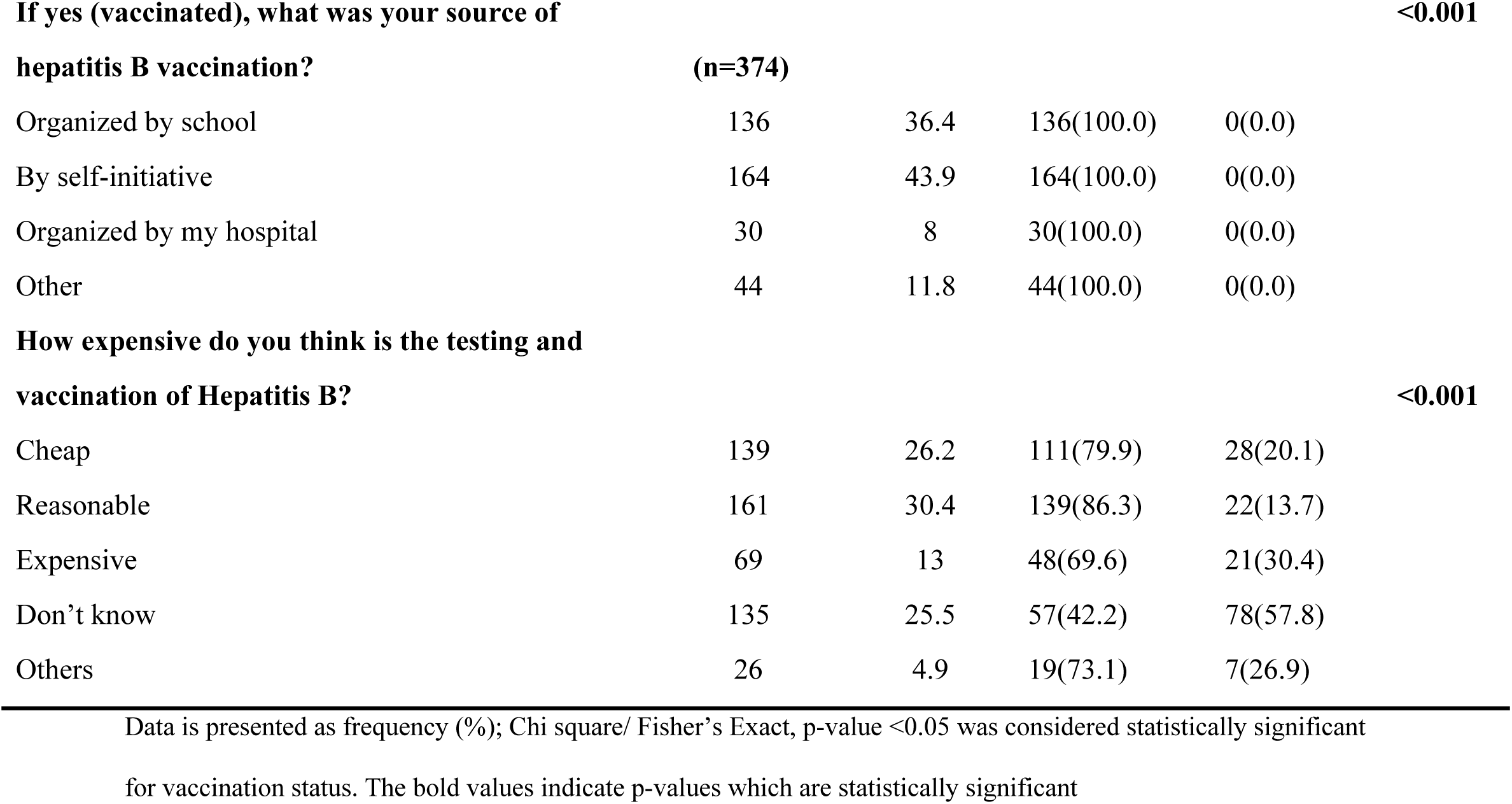
Association between knowledge on Hepatitis B and vaccination status.

| Variable | Frequency | Percentage (%) | Vaccination Status |  | p-value |
| --- | --- | --- | --- | --- | --- |
|  |  |  | Yes (n=374) | No (n=156) |  |
| **Have you done Hepatitis B test Before** |  |  |  |  | **<0.001** |
| Yes | 405 | 76.4 | 355(87.7) | 19(15.2) |  |
| No | 125 | 23.6 | 50(12.3) | 106(84.8) |  |
| **If yes, what was your reason for testing** | **(n=405)** |  |  |  | **<0.001** |
| Just wanted to know my hepatitis B status (Self initiative) | 182 | 44.9 | 156(85.7) | 26(14.3) |  |
| It was an institutional requirement | 125 | 30.9 | 108(86.4) | 17(13.6) |  |
| Testing in order to take the vaccine | 64 | 15.8 | 63(98.4) | 1(1.6) |  |
| Other | 34 | 8.4 | 28(82.4) | 6(17.6) |  |
| **If no, what was your reason for not testing for hepatitis B?** | **(n=125)** |  |  |  | **<0.001** |
| Do not have any reason | 62 | 49.6 | 1(3.4) | 51(82.3) |  |
| Do not get time | 29 | 23.2 | 11(17.7) | 28(96.6) |  |
| Other | 34 | 27.2 | 7(20.6) | 27(79.4) |  |
| **If no, what was your reason for not getting vaccinated (n=156)** |  |  |  |  | **<0.001** |
| Do not have any reason | 70 | 44.9 | 0(0.0) | 70(100.0) |  |
| Busy schedule | 32 | 20.5 | 0(0.0) | 32(100.0) |  |
| Other | 54 | 34.6 | 0(0.0) | 54(100.0) |  |
| **If no, (not vaccinated), will you take the hepatitis B vaccine if it were given free of charge?** | **(n=156)** |  |  |  | **<0.001** |
| Yes | 125 | 80.1 | 0(0.0) | 125(100.0) |  |
| No | 31 | 19.9 | 0(0.0) | 31(100.0) |  |
| **If yes (vaccinated), how many doses of the hepatitis B vaccine have you taken?** | **(n=374)** |  |  |  | **<0.001** |
| 1 | 48 | 12.8 | 48(100.0) | 0(0.0) |  |
| 2 | 95 | 25.4 | 95(100.0) | 0(0.0) |  |
| ≥3 | 231 | 61.8 | 231(100.0) | 0(0.0) |  |

|  |  |  |  |  |
| --- | --- | --- | --- | --- |
| **If yes (vaccinated), what was your source of hepatitis B vaccination?** |  |  |  | **<0.001** |
|  | **(n=374)** |  |  |  |
| Organized by school | 136 | 36.4 | 136(100.0) | 0(0.0) |
| By self-initiative | 164 | 43.9 | 164(100.0) | 0(0.0) |
| Organized by my hospital | 30 | 8 | 30(100.0) | 0(0.0) |
| Other | 44 | 11.8 | 44(100.0) | 0(0.0) |
| **How expensive do you think is the testing and vaccination of Hepatitis B?** |  |  |  | **<0.001** |
| Cheap | 139 | 26.2 | 111(79.9) | 28(20.1) |
| Reasonable | 161 | 30.4 | 139(86.3) | 22(13.7) |
| Expensive | 69 | 13 | 48(69.6) | 21(30.4) |
| Don't know | 135 | 25.5 | 57(42.2) | 78(57.8) |
| Others | 26 | 4.9 | 19(73.1) | 7(26.9) |

Data is presented as frequency (%); Chi square/ Fisher's Exact, p-value <0.05 was considered statistically significant for vaccination status. The bold values indicate p-values which are statistically significant

### Univariate and multivariate logistic regression model of sociodemographic, knowledge on Hepatitis B and vaccination and predictors of vaccination status among study participants

In a univariate logistic regression model, age group, status in hospital, profession, whether or not one has done the Hepatitis B test before, reason for doing or not doing the Hepatitis B test, expensive nature of Hepatitis B testing and vaccination were predictors of vaccination status.

After adjusting for age in the multivariate logistic regression model, have not done hepatitis B test before [aOR= 170.937. 95% CI (19.003-1537.625), p ˂ 0.001], institutional requirement as the reason for testing [aOR= 9.277. 95% CI (1.174-73.323), p=0.035], just wanted to know my hepatitis B status (Self initiative) as the reason for testing [aOR= 8.785. 95% CI (1.143-67.556), p=0.037], other as the reason for testing [aOR= 15.479. 95% CI (1.715-139.670), p=0.015], and don’t know how expensive Hepatitis B testing and Vaccination is [aOR= 2.993. 95% CI (1.392-6.435), p=0.005] were the independent predictors of Hepatitis B vaccination status (**Table 3.0**).

**Table 3:** Univariate and multivariate logistic regression model of sociodemographic, knowledge on Hepatitis B and vaccination and predictors of vaccination status among study participants.

| Variable | cOR (95% CI) | *p*-value | aOR (95% CI) | *p*-value |
| --- | --- | --- | --- | --- |
| **Age Group (In Years)** |  |  |  |  |
| <20 | 0.428(0.154-1.186) | 0.103 | 1.184(0.191-7.324) | 0.856 |
| 20-30 | 0.941(0.445-1.987) | 0.872 | 0.605(0.152-2.415) | 0.477 |
| 31-40 | 3.056(1.247-7.490) | **0.015** | 0.687(0.213-2.221) | 0.531 |
| >40 | 1.00 |  |  |  |
| **Marital Status** |  |  |  |  |
| Single | 1.00 |  |  |  |
| Married | 5.261(0.318-87.159) | 0.246 | - | - |
| Divorced | 1.909(0.118-30.766) | 0.648 | - | - |
| **Profession** |  |  |  |  |
| Clinician | 0.595(0.205-1.723) | 0.338 | 0.882(0.209-3.711) | 0.864 |
| Nurse/Midwife | 0.683(0.318-1.469) | 0.329 | 0.516(0.183-1.454) | 0.211 |
| Others | 0.243(0.104-0.573) | **0.001** | 2.333(0.760-7.158) | 0.139 |
| Pharmacist | 0.527(0.157-1.769) | 0.3 | 0.624(0.118-3.283) | 0.557 |
| Lab Scientist | 1.00 |  |  |  |
| **Status In Hospital** |  |  |  |  |
| Non-Permanent Staff | 1.00 |  |  |  |
| Permanent Staff | 2.574(1.754-3.777) | **<0.001** | 0.694(0.356-1.353) | 0.283 |
| **Have you done Hepatitis B test Before** |  |  |  |  |
| Yes | 1.00 |  |  |  |
| No | 39.611(22.378-70.114) | **<0.001** | 170.937(19.003-1537.625) | **<0.001** |
| **If yes, what was your reason for testing** |  |  |  |  |
| It was an institutional requirement | 0.101(0.013-0.776) | **0.028** | 9.277(1.174-73.323) | **0.035** |
| Just wanted to know my hepatitis B status (Self initiative) | 0.095(0.013-0.717) | **0.022** | 8.785(1.143-67.556) | **0.037** |
| Other | 0.074(0.009-0.644) | **0.018** | 15.479(1.715-139.670) | **0.015** |
| Testing in order to take the vaccine | 1.00 |  |  |  |
| **If no, what was your reason for not testing for hepatitis B?** |  |  |  |  |
| Do not have any reason | 198.8(26.464-1493.395) | **<0.001** | 0.959(0.313-2.939) | 0.942 |
| Do not get time | 6.039(0.741-49.237) | **0.093** | 8.912(0.989-80.315) | 0.051 |
| Other | 1.00 |  |  |  |
| **How expensive do you think is the testing and vaccination of Hepatitis B?** |  |  |  |  |
| Cheap | 0.627(0.340-1.157) | 0.135 | 1.160(0.535-2.518) | 0.707 |
| Don't know | 0.116(0.066-0.203) | **<0.001** | 2.993(1.392-6.435) | **0.005** |
| Expensive | 0.362(0.183-0.716) | **0.003** | 1.408(0.558-3.555) | 0.469 |
| Others | 0.430(0.162-1.140) | 0.09 | 1.935(0.497-7.534) | 0.341 |
| Reasonable | 1.00 |  |  |  |

cOR, Crude Odd ratio; aOR, Adjusted Odd ratio; CI, Confidence interval; 1.00, reference. Binary logistic regression
analysis performed to obtain odd ratios. p-value of < 0.05 was considered statistically significant. The bold values
indicate p-values which are statistically significant

## Discussion

Ghana is one of the countries in Africa classified as being highly endemic to HBV infection [24]. The national HBV prevalence is higher than 8%, which means that everyone living in Ghana is at a high risk of contracting HBV infection [10,12]. As a results of the nature of their work, HCWs in this country even have a higher risk of contraction HBV infection compared to the general Ghanaian population. This makes it relevant for all HCWs in Ghana to adhere to the complete HBV vaccination schedule [25–28].

Even though 70.6% of study participants self-reported having taken at least one dose of the hepatitis B vaccine (≥ 1 dose) with 29.4% having taken no dose of the vaccine at all, the overall complete vaccination coverage (≥ 3 doses) was low (43.6%). This seemingly low complete vaccination coverage (43.6%) is comparable to similar studies (42.3%, 46.8%) in HCWs in other regions of Ghana [29, 30]. It is also consistent with findings from another study by Issa et al. [31] (42.0%) in Nigeria. This vaccination coverage is far lower than what is expected of a high-risk group like HWCs per the recommendation by WHO and Centre for Disease Control and Prevention (CDC) [18, 32]. Still, finding from this current study is higher than what was observed by other studies in Africa. For instance, complete vaccination coverage (≥ 3 doses) was reported by Biset AM & Adugna HB, 2017 to be 28.7% in Ethiopia [33], 24.5% in Cameroon by Noubiap *et al*., 2014 [34], 16.4% in Somalia by Hussein *et al.* [35], 12.9% in Ethiopia by Abebaw et al. [36] and 10.9% in Burkina Faso by Ouédraogo *et al.* [37]. This brings to the fore, the generally low vaccination coverage in Ghana and African at large [38,39]. The observed complete vaccination coverage in this study is lower compared to what has been found in a study by Yuan *et al.* (60%) in China [40] and Guthmann *et al.* (91.7%) in France [41].

The study found that 76.4% of participants had undergone Hepatitis B testing at least once in their lifetime. Since the hepatitis B testing is prerequisite for the vaccination, it is suggestive of potential likelihood of vaccination compliance and the high awareness of the risk of HBV infection on the part of the participants. Significant associations were found between demographic factors such as age, marital status, professional status and vaccination status. Specifically, in the present study, participants aged 31-40 years had the highest (≥ 1 dose) vaccination rate (87.0%), with the least among those aged below 20 years (48.3%). This is not surprising as literature supports the notion that individuals older than 20 years may have higher vaccination rates due to longer exposure to healthcare settings and potentially greater awareness of the importance of vaccination [16]. Also, congruent with findings of this research, a study conducted in Ghana among university students had majority of the vaccinees to be 26 years and above [42]. Meanwhile, previous research has also shown that younger age groups tend to have higher vaccination rates, possibly due to increased awareness and education [19]. This is relative as the younger could be in line with the 31-40 years age group.

The major motivating factor for testing was to know their Hepatitis B infection status. This is in line with existing literature, emphasizing the importance of individuals’ motivation to know their health status, particularly regarding infectious diseases like Hepatitis B [17]. Self-initiative being the commonest impulse of Hepatitis B vaccination is suggestive that proactive behaviour among healthcare workers plays a crucial role in ensuring vaccination uptake [14]. According to a related study, the common reasons given for not getting vaccinated were the non-obligatory nature of the Hepatitis B vaccination, lack of knowledge about the shot, its expensive cost, lack of interest or desire and problems with availability [16, 29].

While the association between sex and vaccination status was not statistically significant, there was a significant association between marital status and vaccination status (p < 0.001). Married individuals had a higher vaccination rate compared to single individuals. These findings suggest that marital status may influence vaccination behaviour, possibly due to shared healthcare decisions among married couples [14]. This could also be attributed to the pre-marital laboratory screening testing for would-be couples. A study carried out among Nigerians; higher rate of hepatitis B vaccinations were observed among married couples [43]. This underscores the importance of tailored vaccination strategies targeting different age groups and marital statuses. The associations between region, religion, and educational level with vaccination status were not statistically significant. While these factors did not show significant associations in this study, variations in vaccination uptake across regions, religious beliefs, and educational levels have been reported in other settings and therefore warrant further investigation [10].

Regardless of health facility, there was a significant association between profession and status in the hospital with vaccination status [35, 44]. These findings highlight the influence of occupational factors and employment status on vaccination behaviour among healthcare workers, with clinical staff members and permanent employees showing higher vaccination rates [14]. This is possible due to the fact that, clinical and permanent staff members are often taken through routine health assessments prior to their posting to their working stations. According to a Ghanaian study consistent with the proposition of the current study findings, working for more than 16 years, doing invasive procedures routinely, being around blood-stained linens and garbage, and being exposed to blood and or its products on a daily basis were all linked to higher hepatitis B vaccination rates [29].

The finding that participants who had undergone Hepatitis B testing had significantly higher vaccination rates aligns with previous research where individuals who are aware of their Hepatitis B status are more likely to seek vaccination if they are unvaccinated [16]. In a study involving 114 undergraduate students studying public health in Ghana, about half (50.4%) had never had their HBV infection checked, and 100 (44.2%) had at least one dose of the vaccine [45]. This implies that, the greater the number of individuals participating in pre-vaccination hepatitis B screening exercise, the greater the fraction of potential coverage of the vaccination. The present study recording the primary reason for testing being self-initiative reflects a proactive approach toward health awareness among the study participants. This is consistent with findings from studies emphasizing the importance of individual motivation and health-seeking behaviour in infectious disease prevention [14,46]. Relatively high number participants (43.9%) reported to have taken the vaccine as a result of their own initiative and good proportion (36.4%) also reported having been given the vaccine by their schools. Only a small proportion (8%) reported having taken the vaccine as a result of vaccination exercise organized by their workplace (hospitals). This area needs attention since schools and workplaces arranging for hepatitis B vaccinations for its students and workers respectively could increase vaccination coverage in Ghana. In this current study, most (80.1%) of the unvaccinated respondents were ready to take the vaccine if given at no cost. Efforts to reduce or eliminate costs associated with testing and vaccination could potentially improve vaccination coverage among healthcare workers.

Interestingly, among those who had not received any dose of the vaccine, a significant proportion (44.9%) mentioned that they did not have any reason for not getting vaccinated. This suggests that occupational health and safety programmes, infection prevention and control programmes, together with health promotion campaigns in the various health facilities could reiterate the need for vaccination adherence and subsequent improved vaccination coverage. The current study also found that, only 13% of the participants reported that the cost of HBV vaccination in the country is expensive, while 82.1% of the participants reported that the cost was cheap, expensive or do not know. This suggests also that, the cost of test and vaccination may not be a key reason for low vaccination coverage among this study population. Perhaps increasing awareness and establishment of vaccination policies in health institutions could increase vaccination coverage.

Interestingly, there was a statistically significant differences among professions as far as the vaccination status (≥ 1 dose) is concerned. While laboratory scientists recorded the highest vaccination coverage (80.4%), followed by Nurses/Midwives (73.7%), Clinicians (71.0%), then pharmacists (68.4%), participants from other professions (such as cleaners, administrative staff, etc) recorded the least vaccination coverage (50.0). Meanwhile, a systematic review on HBV vaccination coverage in Africa found clinicians to be having the highest vaccination coverage [38].

### Limitations and strengths of the study

One of the limitations of this study is the possible recall bias on the part of the participants since vaccination status was self-reported and not verified from properly documented sources. Meanwhile, unlike childhood vaccination which could be more difficult to recall, HCWs are less likely to forget vaccination in their adulthood.

This study has several strengths, it has brought to the fore, the current HBV vaccination status of HCWs in Kumasi. Also, it has been able to identify several factors associated with HBV vaccination coverage among HCWs. Furthermore, this study was a multi-centre research making the findings more generalizable, unlike other studies done in the country which involved only one study centre. Again, all cadres of HCWs together with non-medical hospital staff members were included in this study irrespective of the duration of employment or nature of work. In this way, the findings from the current study give a better representation of all HWCs in the facilities selected.

## Conclusion

The hepatitis B vaccination coverage among HCWs as well as other hospital staff members in Kumasi is low. The HBV infection poses a significant public health challenge in a country like Ghana which has been classified as endemic to HBV infection. We therefore propose implementing programmes for infection prevention and control, occupational health and safety, and health promotion campaigns across the healthcare facilities in Ghana to improve vaccination coverage. Additionally, we recommend conducting hepatitis B screening and vaccination initiatives in all tertiary academic institutions, and employers in Ghana should arrange these services for their employees including the temporary and non-medical staff members.

## Data Availability

The data can be accessed online via https://doi.org/10.5061/dryad.tmpg4f56b

## Acknowledgements

We are much grateful to the management of all the hospitals involved in this study. We also express our appreciation to all individuals who willingly participated in the study.

